# Development of a mobile laboratory system in hydrogen fuel cell buses and evaluation of the performance for COVID-19

**DOI:** 10.1101/2023.01.30.23285220

**Authors:** Miho Okude, Kenji Suzuki, Asami Naito, Akio Ebashi, Tomoka Kusama, Junichi Kiyotaki, Yusaku Akashi, Yoshihiko Kiyasu, Yoko Kurihara, Shigeyuki Notake, Masaki Takanashi, Tomokazu Setoyama, Yasushi Kawakami, Hiromichi Suzuki

## Abstract

**Introduction:** We newly designed and developed two types of hydrogen fuel cell (HFC) buses (motorcoach type and minibus type) with a mobile laboratory system. Feasibility studies have been performed for mobile laboratory testing, especially for the laboratory performance of COVID-19 RT-PCR (PCR).

**Methods:** We evaluated the driving range capability, PCR sample size capacity, turn-around time (TAT), and analytical performance for the detection of SARS-CoV-2. Saliva samples were used for the current research and the analytical performance was compared with reference PCR.

**Results:** The estimated driving range and sample size capacity were 432 km and 3,258 samples, respectively for the HFC motorcoach and 313 km and 2,146 samples for the HFC minibus, respectively. For the TAT, the median time between the sample submission and the completion of PCR were 86 min for the motorcoach and 76 min for the minibus, and the median time between sample submission and the electronic reporting of the result to each visitor were 182 min for the motorcoach and 194 min for the minibus. A secondary analysis of 1,574 HFC mobile laboratory testing samples was conducted and all negative samples were negative by reference PCR. Furthermore, all positive samples were confirmed as positive by reference PCR or other molecular examinations.

**Conclusion:** We confirmed the feasibility of HFC mobile laboratory systems for achieving the rapid reporting of highly accurate PCR results.

## Introduction

Automated laboratory testing has a significant advantage for rapid, accurate and high-throughput laboratory testing. However, the need for heavy medical devices and electronic power are barriers to utilization at sample collection sites; thus, samples must be transported to centralized laboratories for laboratory testing. Consequently, it takes days for patients to obtain their results. Point-of-Care testing (POCT), including rapid antigen testing [1, 2] and POCT type molecular testing [3-7] overcome these problems; however, the sensitivity of antigen testing is inferior to that of molecular testing [2] and POCT-type molecular testing is not suitable for the analysis of many samples.

Mobile laboratory systems are alternative options for laboratory systems to manage a large volume of laboratory samples with rapid reporting [8] and were clinically implemented during the COVID-19 pandemic [9-12]. A hydrogen fuel cell bus (HFC-bus) is a bus that uses hydrogen as a power source. Water and heat are the only by-product of HFC vehicles. An HFC-bus has a large capacity for energy storage and electricity supply, and also has the feature of low vibration amplitude for conversion of hydrogen energy to electric power, both of which are significant merits for a mobile clinical laboratory.

In 2021, we newly designed and developed two types of HFC-buses equipped with a laboratory system for disaster infection control (Figure 1): a motorcoach type HFC-bus (HFC motorcoach) and a minibus type HFC-bus (HFC minibus). The motorcoach was a remodeled SORA (Toyota Motor Corporation), which was the first commercially available HFC-bus in Japan. The HFC minibus was fabricated as a prototype vehicle by Toyota Motor Corporation by retrofitting fuel cell system on a diesel-powered minibus (COASTER; Toyota Motor Corporation) and was remodeled for current project for use as a mobile laboratory system. Both HFC-buses were equipped with automated laboratory systems with electronically reporting, and patients can receive their laboratory results via e-mail soon after the submission of clinical samples.

**Figure 1.**
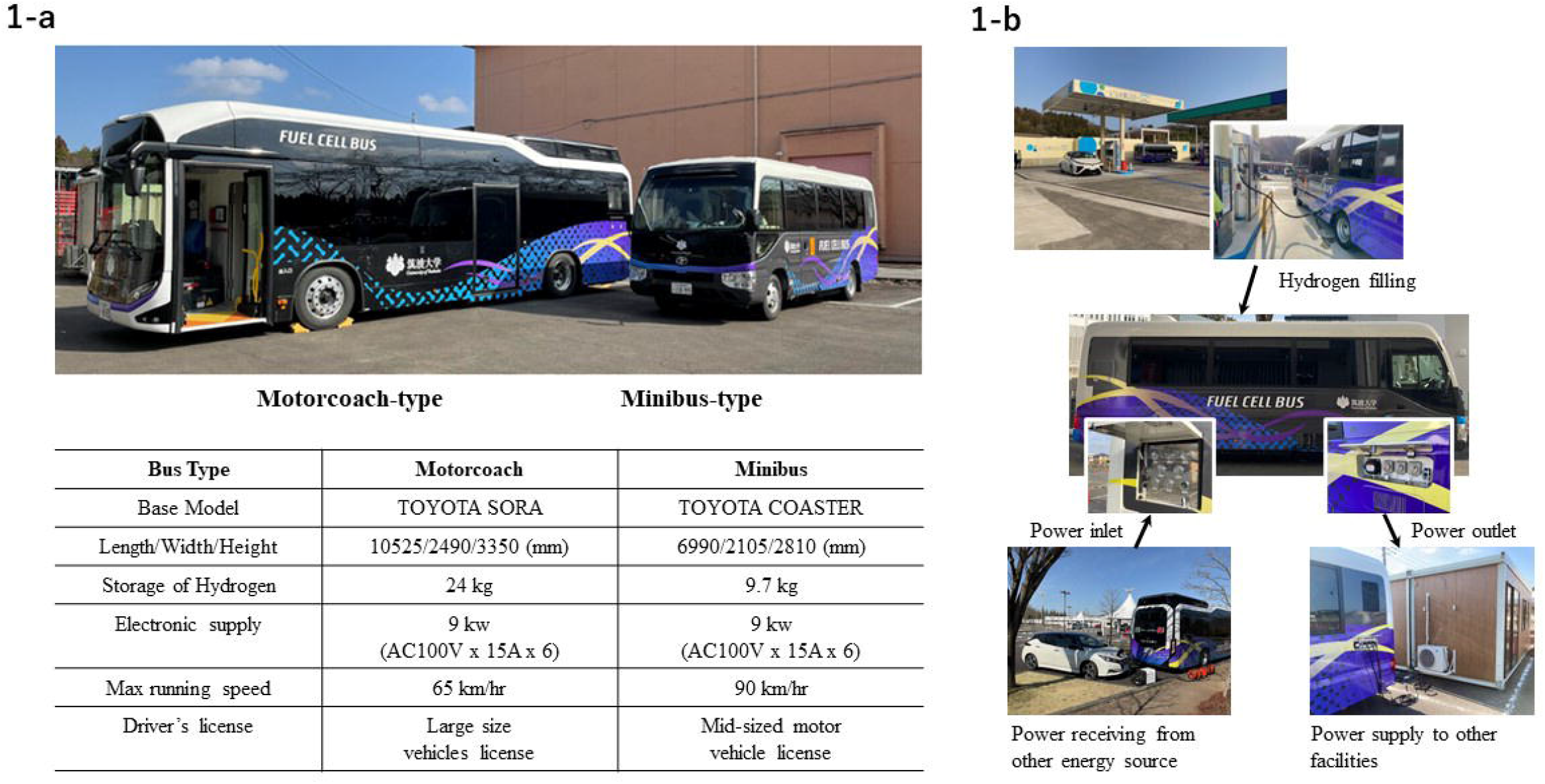
General information about the two hydrogen fuel cell buses (motorcoach type and minibus type). 1-b shows the power supply function and the power receiving function.

In the current study, we evaluated the performance of the mobile laboratory system on the HFC-buses, focusing on RT-PCR for COVID-19. The mobile range, laboratory capacity, turn-around-time (TAT) and accuracy of laboratory testing were evaluated.

## Methods

The current study was performed between October, 2021 and March 2022. The evaluation was performed in Tsukuba City Office, Ibaraki Prefectural Office, public health centers, temporary PCR centers and the University of Tsukuba Hospital. Evaluation of the mobile range was made based on the hydrogen volume consumed by actual driving and RT-PCR testing. TAT was measured between patient sample submission and the reporting of results via e-mail to each device. The accuracy of laboratory testing was evaluated in comparison to a reference RT-PCR assay in a centralized laboratory. The ethics board of the University of Tsukuba Hospital approved the present study (approval number: R03-043) including the study protocol. The requirement for obtaining informed consent was waived for the evaluation of anonymized samples.

### Specifications of the motorcoach-type HFC-bus and the minibus-type HFC-bus

Both buses were designed to achieve a maximum electricity output of 9 kW (1.5kW x 6 outlets, 100V) for electricity for the laboratory (Figure 1-a) and to meet the requirement of biohazard level 2 (P2) according to the WHO criteria. Both buses could supply and receive up to 9 kW of electricity (Figure 1-b). The quality of the power output was confirmed to meet the requirements for medical electrical equipment under ambient temperatures (30°C/0°C). Both buses were certified to run on public roads before the current study. Both buses were permitted only 1 person as a driver and no passenger could ride the bus while it was driven.

The general flow of the mobile laboratory systems is shown in Figure 2. The laboratory system (Figure 2-a) included an electronic reservation system, a data management system, safety cabinets, refrigerators for samples and reagents, automated purification systems (magLEAD, Precision System Science Co., Ltd., Chiba, Japan), automated PCR examination systems (GENECUBE, TOYOBO Co., Ltd., Osaka, Japan), and an electronic reporting system. Reservations can be performed by the patient using an electronic device and the laboratory results are sent to their registered e-mail address soon after the submission of the samples (Figure 2-c). The detailed layout of each bus for laboratory testing is described in Supplementary Figure 1.

**Figure 2.**
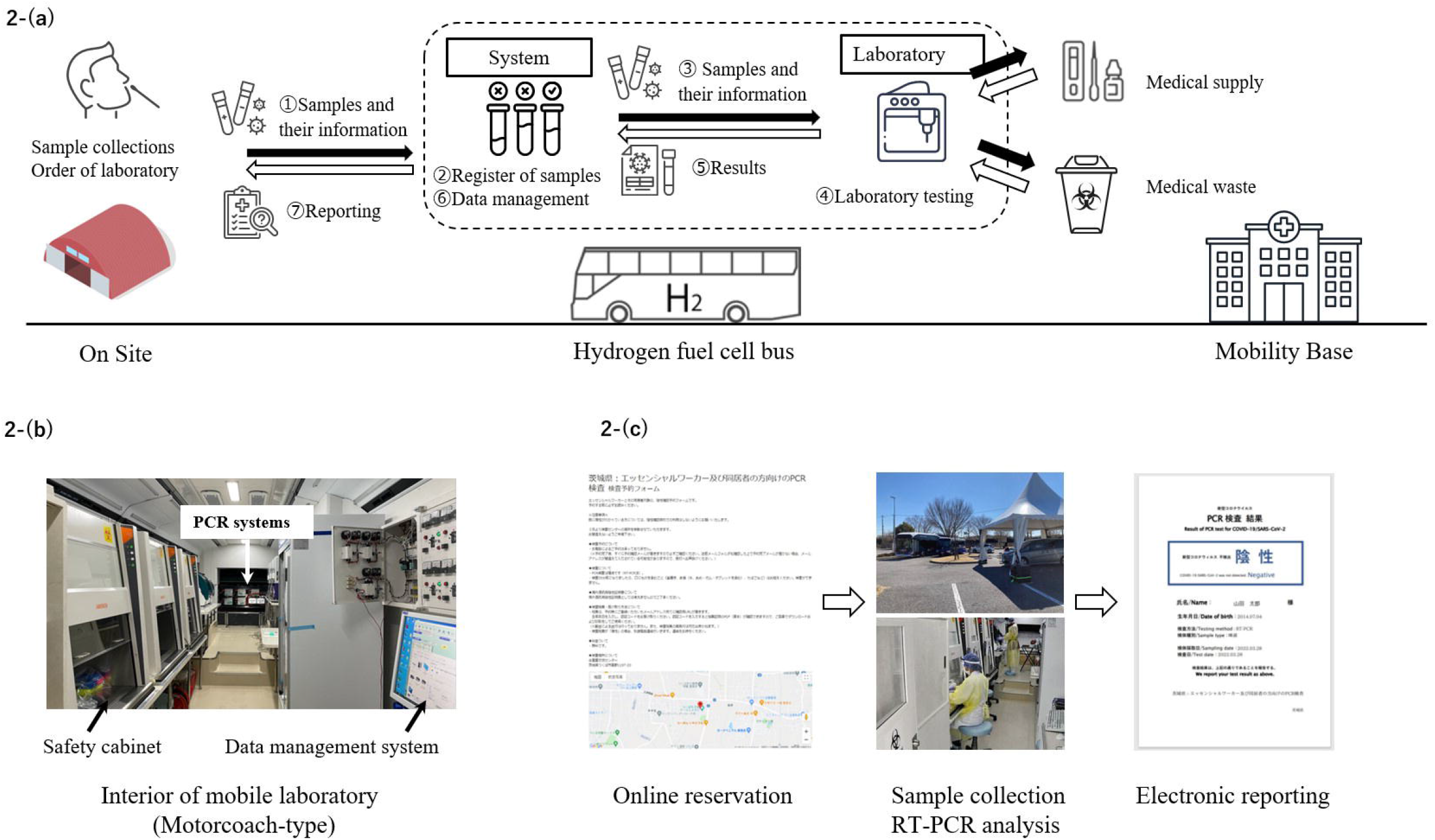
2-(a) shows the flow of the laboratory system in buses, 2-(b) shows a picture of interior of the motorcoach-type hydrogen fuel cell bus and 2-(c) shows the flow of electronic reservation and reporting systems.

GENECUBE is a Qprobe-PCR-based automated rapid molecular identification system that can detect target genes in a short time and simultaneously analyze up to 24 samples and 4 assays in a single examination. The system automatically performs a direct molecular examination, including preparation of the reaction mixtures, and amplification and detection of target genes, in 30 minutes. GENECUBE HQ SARS-CoV-2 (TOYOBO Co., Ltd.) [13], along with rapid purification methods with magLEAD were used to conduct molecular examinations in the buses [14]. Through the examination from preparation of samples to RT-PCR, the entire procedure was performed in the buses using plastic droppers. Positive and negative samples were set to each run in this study, and all positive samples were re-evaluated before reporting.

### Hydrogen fuel consumption for driving and laboratory testing (COVID-19 RT-PCR)

Hydrogen fuel consumption by the motorcoach and minibus was measured during a one way-trip to several different testing sites from the University of Tsukuba Hospital, with RT-PCR examinations conducted in those places. Negative anonymized saliva samples and SARS-CoV-2 reference material; AccuPlex™ SARS-CoV-2 Reference Material Kit (SeraCare; SeraCare Life Sciences, Inc., Milford, MA, USA) were used for laboratory testing.

### Measurement of laboratory testing (COVID-19 RT-PCR) turn-round-time (TAT)

First, we measured 5 saliva samples to evaluate the turn-around-time of RT-PCR from sample submission to reporting the results using the laboratory system. All samples were provided by volunteers who had provided their written informed consent.

Secondly, we measured the TAT between sample submission and reporting to each electronic device (e.g., mobile phones) from December 15, 2021 to December 17, 2021 in Tsukuba City Office PCR center. Tsukuba City has provided RT-PCR testing of saliva samples for asymptomatic residents and commuters in Tsukuba City Office. Participants who had provided their written informed consent provided saliva samples for the TAT analysis. For the second TAT evaluation, pooled-sample PCR screening (5 samples) was used because the city provided a pooled RT-PCR method for visitors. Both the first TAT analysis and the second TAT evaluation were performed only in the motorcoach.

### Comparison of the diagnostic accuracy of laboratory testing (COVID-19 RT-PCR) in the HFC-bus and that of the reference RT-PCR assay in a centralized laboratory

Between February 1, 2022 and March 31, 2022, HFC-buses were sent to two temporary COVID-19 PCR centers in Tsukuba at the request of Ibaraki Prefectural Office (Supplementary Figure 2). Asymptomatic workers at clinic, hospitals or social welfare facilities who were in close contact with COVID-19 patients in Ibaraki Prefecture used the temporary PCR centers. Eligible individuals made an online reservation and submitted samples saliva by the drive-through method in the temporary PCR centers (Figure 2-c). The samples were immediately analyzed in the buses and the results were reported to visitors electronically through their registered email address. We analyzed the data and performed the analysis of the diagnostic accuracy of COVID-19 RT-PCR between February 1, 2022 and February 28, 2022 for the HFC motorcoach and between March 23, 2022 and March 28, 2022 for the HFC minibus. Residual saliva samples were anonymized and preserved at –80°C until the evaluation.

For the analytical evaluation, the preserved frozen saliva samples were sent to LSI Medience Corporation for comparison, then a reference real-time RT-PCR examination was performed using a Maxwell^®^ RSC Viral Total Nucleic Acid Purification Kit for RNA extraction (Promega Corporation, WI, USA) according to the manufacturer’s instructions and a Cobas® Z480 Real-Time PCR System (Roche, Basel, Switzerland) using a method developed by the National Institute of Infectious Diseases (NIID), Japan, for SARS-CoV-2 [15]. A duplicate analysis for N2 genes was performed for the evaluation of SARS-CoV-2. The Ampdirect 2019 Novel Coronavirus Detection Kit (Shimadzu Corporation, Kyoto, Japan) [16] and cobas 8800 system and cobas SARS-CoV-2 & Influenza A/B (cobas; Roche Molecular Systems, Branchburg, NJ, USA) [17] were used to evaluate discrepant samples. The differences in the limit of detection among molecular examinations for COVID-19 are shown in Supplementary Table 1.

### Statistical analyses

The total concordance rate, positive concordance rate, and negative concordance rate were calculated with 95% confidence intervals (CIs). The correlation between the Ct value of the real-time RT-PCR assay (NIID method) and the Sp value of the GENECUBE was evaluated using Spearman’s rank correlation coefficient. P values of < 0.05 were considered to indicate statistical significance. All statistical analyses were conducted using the R 4.1.2 software program (The R Foundation, Vienna, Austria) with the “readxl”, “tidyverse”, and “epiR” packages.

## Results

### Evaluation of hydrogen fuel consumption in driving and laboratory testing (COVID-19 RT-PCR)

The evaluation of hydrogen fuel consumption in driving and laboratory testing is summarized in Figure 3. For the motorcoach, the average fuel consumption in driving and laboratory testing (COVID-19 RT-PCR) was 18.0 km/kg (range; 15.7–20.1 km/kg) and 135.7 samples/kg (range; 88.1–179.8 samples/kg), respectively. For the minibus, the average fuel consumption in driving and laboratory testing (COVID-19 RT-PCR) was 32.3 km/kg (range; 25.8–41.2 km/kg) and 221.2 samples/kg (range; 141.4–282.8 samples/kg), respectively.

**Figure 3.**
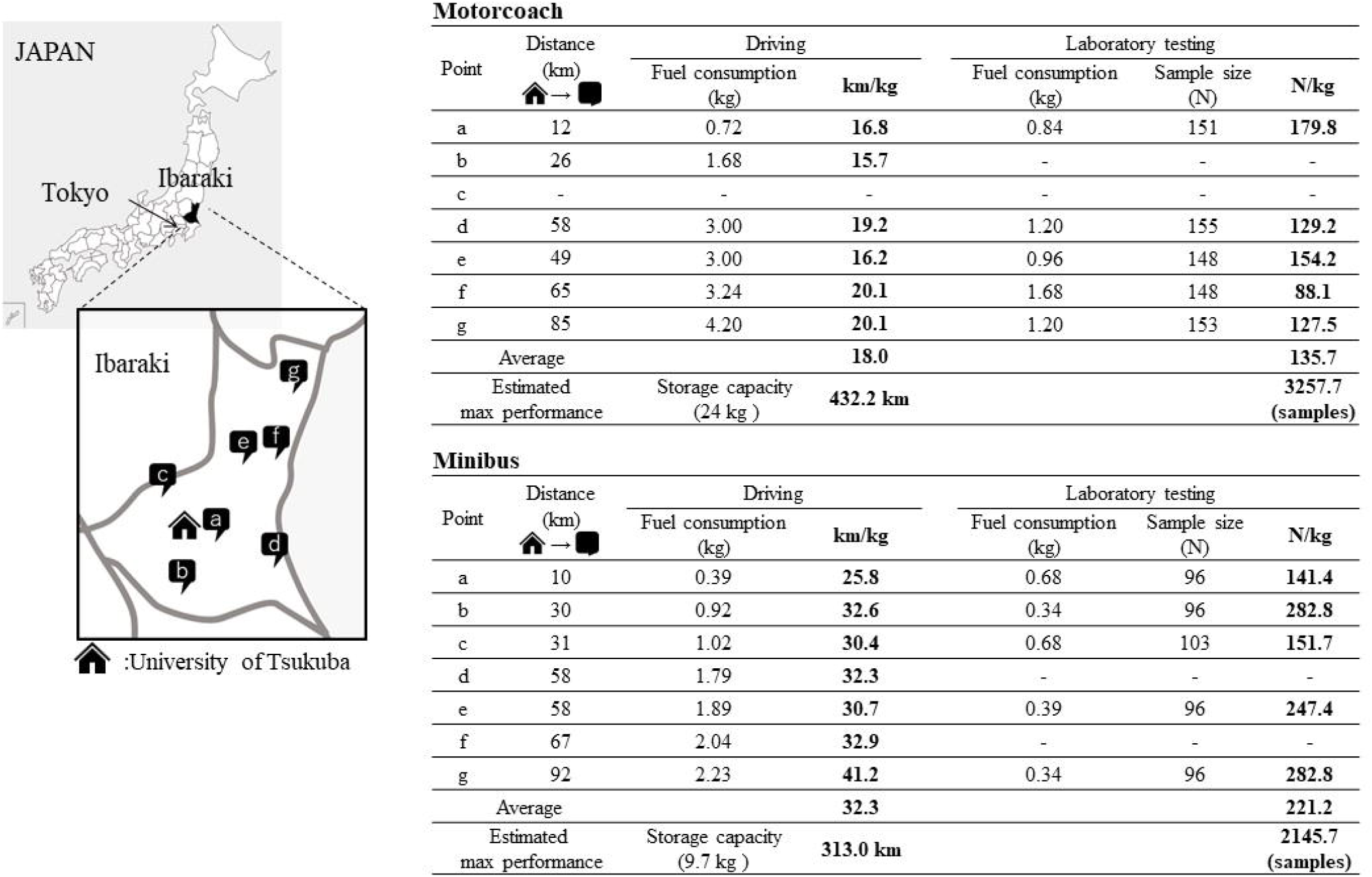
Hydrogen fuel consumption for driving and laboratory testing (COVID-19 PCR). a: Tsuchiura Public Health Center, b: Ryugasaki Public Health Center, c: Chikusei Public Health Center, d: Itako Public Health Center, e: Ibaraki Prefectural Office, f: Hitachinaka Public Health Center, g: Hitachi Public Health Center. Estimated maximum power of driving distance and the laboratory testing throughput of COVID-19 PCR of hydrogen fuel cell bus were calculated according to the hydrogen storage of each bus (motorcoach, 24 kg; minibus, 9.7 kg) and average hydrogen consumption.

### Evaluation of laboratory testing (COVID-19 PCR) turn-around-time (TAT) in the HFC-bus

The first attempt with 5 negative saliva samples showed that the results appeared on the laboratory system 39 minutes after the saliva samples were submitted to the bus.

For the second evaluation at Tsukuba City Office PCR center, written informed consent was obtained from 140 patients and the TAT was evaluated using their samples (Table 1). The median TAT was 54 minutes (IQR; 51-55) and the results were electronically reported in all cases by 59 minutes after submission (Table 1). All samples were found to be negative by RT-PCR.

**Table 1.**
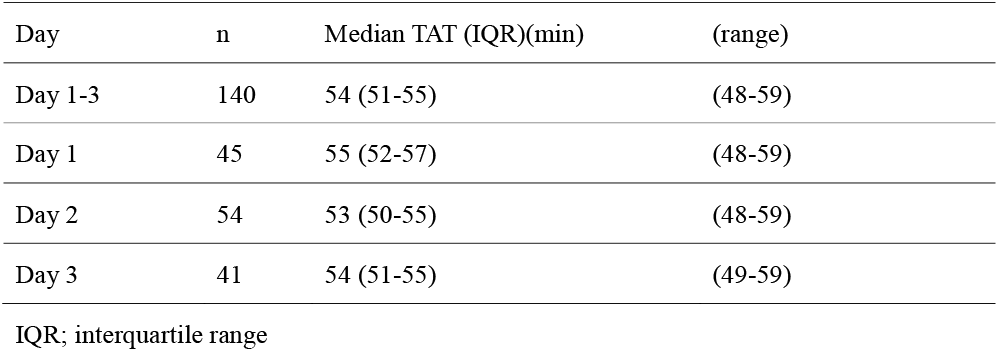
Turn-around time (TAT) between patient sample submission and the reporting of COVID-19 PCR results using a hydrogen fuel cell bus-based mobile laboratory system

### The analysis of the diagnostic accuracy of COVID-19 RT-PCR in the two HFC-buses

During the evaluation period, 18 days of examination data for the motorcoach (n=1395) and 3 days of examination data (n=179) for the minibus were evaluated. The median hydrogen consumption during the COVID-19 RT-PCR was 57.2 samples/kg (IQR; 35.2-90.4) for the motorcoach and 135.0 samples/kg (IQR; 111.8-177.4) for the minibus, respectively (Table 2). For the TAT, the median time between the sample submission and the completion of COVID-19 RT-PCR were 86 min (IQR; 73– 96 min) for the motorcoach and 76 min (IQR; 71-84 min) for the minibus and the median time between sample submission and electronic reporting of the result to each visitor was 182 min (IQR; 134– 272 min) for the motorcoach and 194 min (IQR; 167–227 min) for the minibus, respectively (Table 2). Detailed information was described in Supplementary Table 2.

**Table 2.** Turn-around time (TAT) and hydrogen fuel consumption in COVID-19 PCR centers

|  | Test/day<br>(median) | TAT (IQR) (min)<br>(Sample collection to<br>Patients reporting of<br>the results) | TAT (IQR) (min)<br>(Sample collection to<br>completion of<br>PCR evaluation) | Median<br>fuel consumption<br>(PCR test/kg)<br>(IQR) |
| --- | --- | --- | --- | --- |
| Motorcoach | 61 | 182 (134-272) | 86 (73-96) | 57.2 (35.2-90.4) |
| Minibus | 64 | 194 (167-227) | 76 (71-84) | 135.0 (111.8-177.4) |

IQR; interquartile range

The results of comparison between the molecular examination in buses and a reference RT-PCR assay in the laboratory are shown in Table 3 (n=1574). The total concordance rate, positive concordance rate and negative concordance rate were 98.4% (95%CI: 97.7-99.0%), 100% (95% CI: 95.1–100%) and 98.3% (95% CI: 97.6–98.9%), respectively. Of the 25 bus-positive and reference RT-PCR-negative cases, 23 were positive by an Ampdirect 2019 Novel Coronavirus Detection Kit. The 2 samples were further evaluated with a cobas 8800 system and cobas SARS-CoV-2 & Influenza A/B and all of the two samples were found to be positive.

**Table 3.** Comparison between molecular examinations performed in hydrogen fuel cell buses and the reference RT-PCR assay results in the laboratory

|  |  | real-time RT-PCR in laboratory |  |
| --- | --- | --- | --- |
|  |  | Positive | Negative |
| Molecular examination in bus | Positive | 73 | 25 |
|  | Negative | 0 | 1476 |
| Positive concordance rate (%) |  | 100 (95.1 –100) |  |
| Negative concordance rate (%) |  | 98.3 (97.6- 98.9) |  |

RT-PCR, reverse transcription-polymerase chain reaction
Data in parentheses indicate 95% confidence intervals.

The correlation of the Sp value of GENECUBE in buses and the Ct value of the reference RT-PCR assay was shown in Supplementary Figure 3. The calculation of Spearman’s rank correlation coefficient revealed a significant correlation (Rho = 0.81, p < 0.001) between the Sp value of GENECUBE and the Ct value of the reference RT-PCR assay. The median Sp value was 34.9 (IQR:31.8–38.2) for GENECUBE and the median Ct value was 30.1 (IQR:27.8–31.6) for the reference real-time RT-PCR assay. The median difference was 4.5 (IQR:3.0–5.8).

## Discussion

To date, this is the first report on the clinical implementations of using HFC-buses as mobile laboratories. The current evaluation showed that both types of bus had sufficient hydrogen storage for driving and laboratory testing (COVID-19 PCR). With the implementation of the newly developed mobile laboratory systems in COVID-19 PCR centers, visitors could receive the RT-PCR results within a few hours via online after sample submission. The high diagnostic accuracy for the detection of SARS-CoV-2 RNA in saliva samples was confirmed by comparison with a reference RT-PCR assay.

The clinical implementation of mobile laboratories has dramatically progressed since the onset of the COVID-19 pandemic. In Japan, the clinical use of mobile laboratories was tentatively approved—as a special case—for the COVID-19 pandemic in early February 2022 [18]. This approval allowed the current HFC mobile laboratory systems to be introduced into clinical practice. Touron et al. reported the utility of the Mobil’DNA lab for COVID-19 [11] in French; this transportable laboratory system had been available since 2015. The system used an Applied™ 7500 real-time PCR system operated by highly skilled medical technologists and it was utilized in the COVID-19 pandemic as a high throughput mobile laboratory, which reported results in one day. Ballard also reported the achievement of rapid reporting of molecular examinations by a mobile laboratory system with an automated molecular identification system (GeneXpert) in Australia [9]. They showed that the median TAT from sample collection to reporting of results was approximately 2 hours, which was significantly faster than a conventional laboratory; however, it was limited in the number of samples and the TAT was more than 4 hours in some cases.

In our current HFC-bus-based mobile laboratory system, the laboratory examinations were completed in approximately one hour, and most of results were reported within 3 hours after the samples were obtained, and the TAT was improved to 2 hours (Supplementary Table 3) after adjusting for the workflow in late February. In the 2021-2022 winter season in Japan, medical facilities had to report clinical information of all of newly identified COVID-19 cases, including the patient’s health status, to public health centers. This preparation delayed the TAT in spite of rapid completion of laboratory testing. In the current investigation, the HFC-bus could process one hundred samples in temporary COVID-19 PCR centers without a delay of TAT or a hydrogen energy shortage. In addition, the current system had sufficient analytical performance for the detection of SARS-CoV-2 RNA. Also, laboratory process was performed with automation including reservation and reporting. All of process was easy to handle without pipette during laboratory examination and the current system can be managed by inexperienced technicians in case of emergency use. The current systems required large amounts of electricity for automated laboratory systems and air conditioning (nearly 9 kW) for the appropriate working environment of technicians, and the HFC-bus was essential to supply electricity in addition to the merit of the low vibration amplitude.

Note that the system is not only capable of stand-alone, self-sustaining testing in a HFC-bus, but also has the ability to be connected to other energy resources. Future design implications of such mobile laboratories include the ability to supply power to external facilities and to receive power from an external source, including mobile one such as an EV or Fuel Cell Vehicle (FCV). In this experiment, when weather conditions made it difficult to travel to the hydrogen station, power could be supplied from a FCV during the inspection, and the inspection could then be performed successfully. It is also very important to be able to supply electricity to surrounding environment, such as a small workspace for sample collection and lighting facilities during night. Thus, we believe that a new fleet management will be necessary that not only considers the inspected unit?, but also the whole laboratory system. We consider that we can connect the developed mobile laboratory to medical institutions and public facilities with safe, robust, and stable mobile testing.

The present study was associated with several limitations. First, the hydrogen fuel consumption in driving and laboratory testing was measured just after the development of systems and inexperience may have affected the data. Additionally, the current data were mainly collected in the winter season, which required heating inside the buses; this requirement differs in other seasons. Second, the evaluation of mobile laboratory systems as COVID-19 PCR centers mas mainly performed using the motorcoach type HFC-bus and the data of the minibus type HFC-bus were insufficient. Third, we could not use fresh positive samples for comparison with the reference RT-PCR assay. The storage process involving freezing and thawing is also reported to affect the viral load in samples [19].

In conclusion, the evaluation of the newly developed mobile laboratory system confirmed that HFC-buses have sufficient energy storage for driving and laboratory testing (COVID-19 PCR) and that they are feasible for the rapid reporting of highly accurate COVID-19 RT-PCR results.

## Data Availability

All data produced in the present work are contained in the manuscript.

## Acknowledgements

The current research was funded by the Cross-ministerial Strategic Innovation Promotion Program (SIP, Cabinet Office). We thank Dr. Koki Kaku for his instruction and advice as a subprogram director of the current SIP project. We also thank Mr. Shimpei Miura, Mr. Katsutoshi Tanei and their colleagues (Toyota Motor Corporation) for their technical support and assistance in the development of mobile laboratory system for hydrogen fuel cell buses, Mr. Miyazaki his colleagues (Izumi Motor Car Co., Ltd.) for remodeling the hydrogen fuel cell buses to support the laboratory function, Dr. Yusaku Akashi for support in the statistical analysis, staff of Kanto Railway Co., Ltd. for operating the hydrogen fuel cell buses, city officials of Tsukuba City and the Prefectural Office staff of Ibaraki Prefecture for supporting examination, staffs of ENEOS Corporation for support regarding hydrogen filling, Dr. Toru Nanmoku, Ms. Rikako Kano, and the staff of the University of Tsukuba Hospital, the staff of LSI Medience Corporation, the staff of the National Research Institute for Earth Science and Disaster Resilience for their support of this study.

## Author contributions

Miho Okude performed laboratory testing and drafted the manuscript. Kenji Suzuki drafted the manuscript, created the mobile laboratory system, designed the study and supervised the project. Akio Ebashi provided technical support for the hydrogen fuel cell buses and drafted the manuscript. Asami Naito, Tomoka Kusama, Junichi Kiyotaki performed laboratory testing. Yusaku.Akashi performed the statistical analysis and revised the manuscript. Shigeyuki Notake collected samples, Yoshihiko Kiyasu and Yoko Kurihara revised the manuscript. Masaki Takanashi performed laboratory testing and supervised the project. Tomokazu Setoyama created mobile laboratory system and designed the study. Yasushi Kawakami supervised the project. Hiromichi Suzuki drafted the manuscript, created the mobile laboratory system, designed the study and supervised the project. All authors contributed to the writing of the final manuscript.

## Supplemental Files

Supplementary table 1 The differences in the limit of detection among molecular examinations for COVID-19.

Supplementary Table 2-a data of motorcoach

Supplementary Table 2-b data of minibus

Supplementary Figure 1 Detailed layout of the motorcoach type hydrogen fuel cell bus and minibus type hydrogen fuel cell bus used for laboratory testing.

Supplementary Figure 2 Pictures of two temporary COVID-19 PCR centers, in which hydrogen fuel cell buses were sent.

Supplementary Figure 3 Comparison between the Sp value of GENECUBE and cycle threshold (Ct) values of reference real-time RT-PCR assays (N2 gene).

