## Supplementary files for "Development of a mobile laboratory system in hydrogen fuel cell buses and evaluation of the performance for COVID-19"

Supplementary table 1. The differences in the limit of detection among molecular examinations for COVID-19.

| Copies/mL | Sample | GENECUBE with magLEAD | RT-PCR (N2 NIID method)  with Maxwell | Ampdirect 2019 Novel Coronavirus Detection Kit |
| --- | --- | --- | --- | --- |
|  |  | Detection rate (N of detection) | | |
| 2500 | Total | 100% (40/40) | 97.5% (39/40) | 100% (40/40) |
|  | UTM | 100% (8/8) | 100% (8/8) | 100% (8/8) |
|  | Pooled NP samples | 100% (16/16) | 93.8% (15/16) | 100% (16/16) |
|  | Pooled saliva samples | 100% (16/16) | 100% (16/16) | 100% (16/16) |
| 1000 | Total | 100% (40/40) | 70% (28/40) | 100% (40/40) |
|  | UTM | 100% (8/8) | 87.5% (7/8) | 100% (8/8) |
|  | Pooled NP samples | 100% (16/16) | 81.3% (13/16) | 100% (16/16) |
|  | Pooled saliva samples | 100% (16/16) | 50% (8/16) | 100% (16/16) |
| 500 | Total | 92.5% (37/40) | 15% (6/40) | 90% (36/40) |
|  | UTM | 100% (8/8) | 25% (2/8) | 100% (8/8) |
|  | Pooled NP samples | 93.8% (15/16) | 18.8% (3/16) | 87.5% (14/16) |
|  | Pooled saliva samples | 87.5% (14/16) | 6.3% (1/16) | 87.5% (14/16) |
| 250 | Total | 45% (18/40) | 15% (6/40) | 67.5% (27/40) |
|  | UTM | 62.5% (5/8) | 12.5% (1/8) | 75% (6/8) |
|  | Pooled NP samples | 37.5% (6/16) | 25% (4/16) | 50% (8/16) |
|  | Pooled saliva samples | 43.8% (7/16) | 6.3% (1/16) | 81.3% (13/16) |
| 0 | Total | 0% (0/40) | 0% (0/40) | 0% (0/40) |
|  | UTM | 0% (0/8) | 0% (0/8) | 0% (0/8) |
|  | Pooled NP samples | 0% (0/16) | 0% (0/16) | 0% (0/16) |
|  | Pooled saliva samples | 0% (0/16) | 0% (0/16) | 0% (0/16) |

NP, Nasopharyngeal,

SARS-CoV-2 reference material (AccuPlex™ SARS-CoV-2 Reference Material Kit, SeraCare; SeraCare Life Sciences, Inc., Milford, MA, USA) was diluted with matrix (two UTM™; four pooled negative nasopharyngeal samples and four pooled negative saliva samples) to make 5 difference concentration of samples. The molecular examination with each assay was performed four times for each sample.

Supplementary Table 2. Turn-around time (TAT) and hydrogen fuel consumption of motorcoach type hydrogen fuel cell bus and minibus type hydrogen fuel cell bus

| Table 2-a; data of motorcoach | | | | | |
| --- | --- | --- | --- | --- | --- |
| Date | Sample size (N) | Median TAT (IQR) (min)  (Sample collection to patients reporting of the results) | Median TAT (IQR) (min)  (Sample collection to completion of PCR evaluation) | Fuel consumption (kg) | N/kg |
| 2022/2/1 | 28 | 100 (84-116) | 65 (61-65) | 1.44 | 19.4 |
| 2022/2/2 | 21 | 134 (133-140) | 62 (62-63) | 1.68 | 12.5 |
| 2022/2/3 | 25 | 135 (110-136) | 54 (51-66) | 0.96 | 26.0 |
| 2022/2/4 | 26 | 144 (80-172) | 65 (60-77) | 0.84 | 31.0 |
| 2022/2/7 | 125 | 220 (186-311) | 89 (81-97) | 1.56 | 80.1 |
| 2022/2/8 | 124 | 255 (186-296) | 89 (83-96) | 1.8 | 68.9 |
| 2022/2/9 | 49 | 270 (161-279) | 77 (58-92) | - | - |
| 2022/2/10 | 59 | 290 (156-302) | 85 (66-89) | - | - |
| 2022/2/14 | 169 | 250 (225-344) | 112 (101-116) | 1.56 | 108.3 |
| 2022/2/15 | 53 | 255 (246-266) | 80 (74-92) | 1.2 | 44.2 |
| 2022/2/16 | 44 | 228 (133-257) | 72 (62-89) | 1.2 | 36.7 |
| 2022/2/17 | 63 | 275 (238-290) | 79 (72-84) | 1.32 | 47.7 |
| 2022/2/18 | 81 | 289 (195-312) | 80 (69-89) | 0.84 | 96.4 |
| 2022/2/21 | 144 | 149 (128-167) | 82 (75-92) | 1.32 | 109.1 |
| 2022/2/22 | 85 | 127 (101-139) | 90 (84-96) | 1.32 | 64.4 |
| 2022/2/24 | 54 | 184 (168-199) | 59 (58-79) | 1.08 | 50.0 |
| 2022/2/25 | 106 | 133 (118-156) | 82 (76-95) | 1.2 | 88.3 |
| 2022/2/28 | 139 | 135 (118-160) | 89 (80-95) | 1.32 | 105.3 |

| Table 2-b; data of minibus | | | | | |
| --- | --- | --- | --- | --- | --- |
| Date | Sample size (N) | Median TAT (IQR)  (min)  (Sample collection to patients reporting of the results) | Median TAT (IQR) (min)  (Sample collection to completion of PCR evaluation) | Fuel consumption (kg) | N/kg |
| 2022/3/23 | 64 | 229 (211-249) | 82 (68-93) | 0.29 | 219.9 |
| 2022/3/24 | 43 | 169 (156-190) | 72 (71-84) | 0.49 | 88.7 |
| 2022/3/28 | 72 | 175 (147-216) | 76 (71-84) | 0.53 | 135.0 |

IQR; interquartile range

Supplementary Figure 1. Detailed layout of the motorcoach type hydrogen fuel cell bus and minibus type hydrogen fuel cell bus used for laboratory testing.

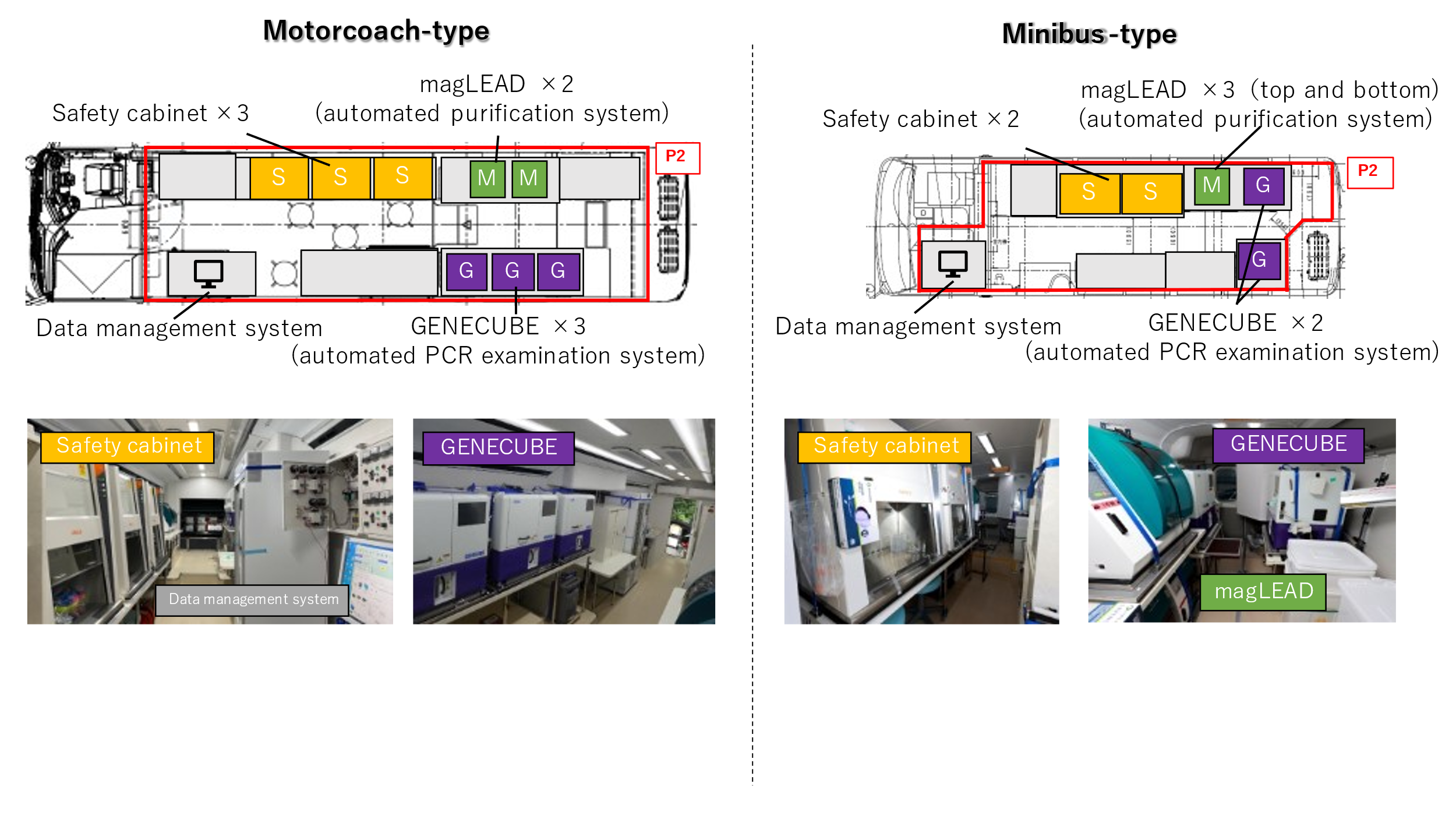

Supplementary Figure 2. Pictures of two temporary COVID-19 PCR centers, in which hydrogen fuel cell buses were sent.

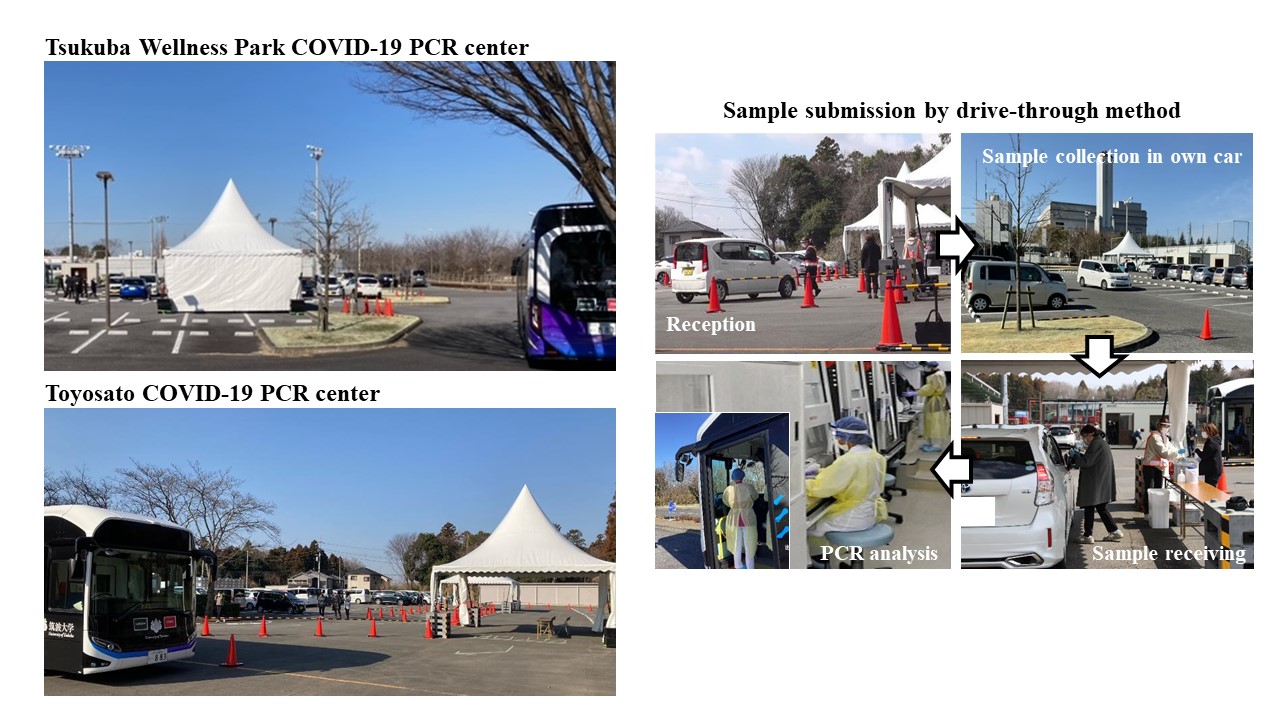

Supplementary Figure 3. Comparison between the Sp value of GENECUBE and cycle threshold (Ct) values of reference real-time RT-PCR assays (N2 gene). Spearman's rank correlation coefficient (R) between the two tests was 0.81. The 25 GENECUBE-positive and reference RT-PCR-negative cases are not included.

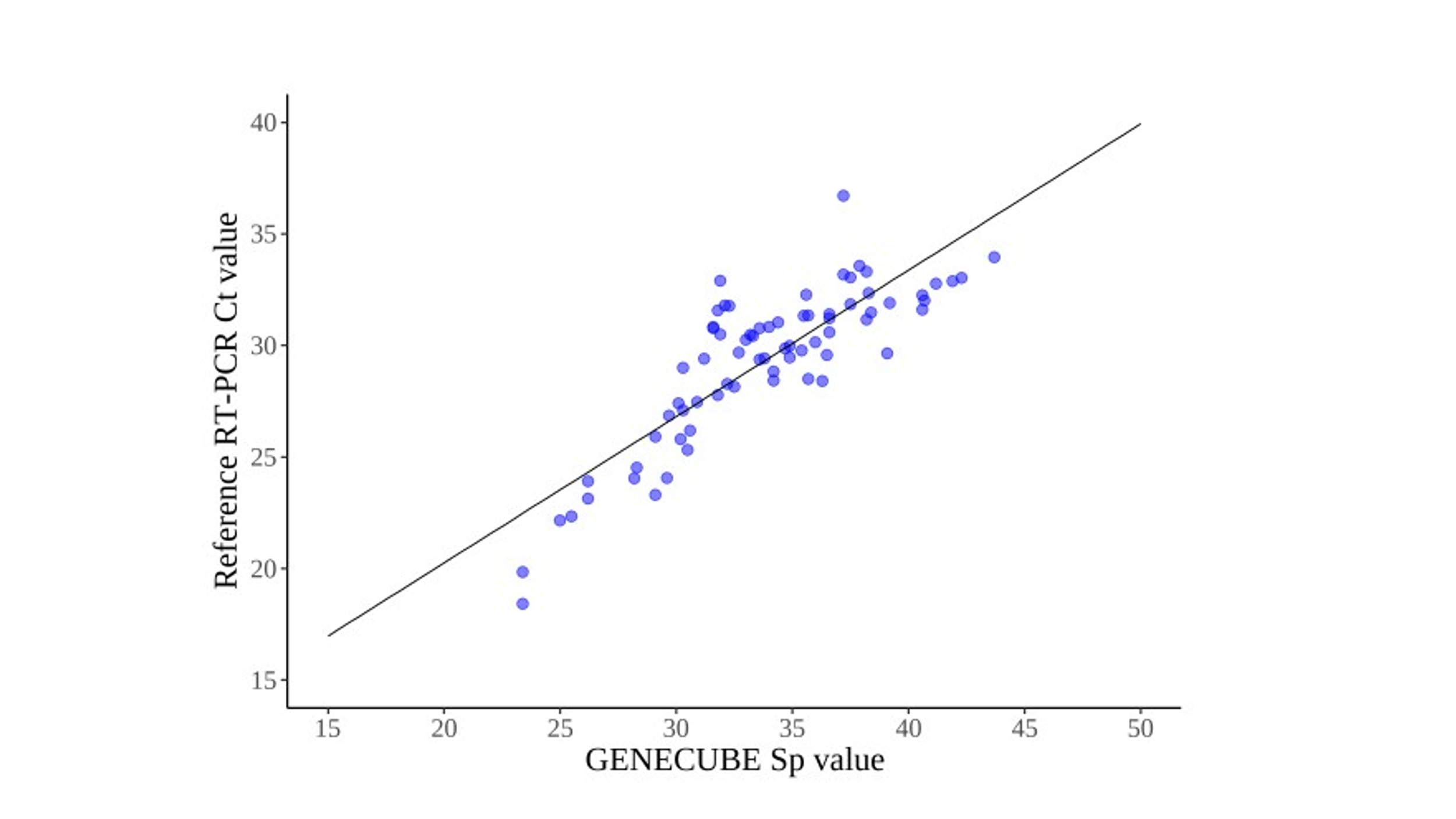
